# Real-time IP-10 measurements as a new tool for inflammation regulation within a clinical decision support protocol for managing severe COVID-19 patients

**DOI:** 10.1101/2020.07.21.20158782

**Authors:** Shaul Lev, Tamar Gottesman, Gal Sahaf Levin, Doron Lederfein, Evgeny Berkov, Dror Diker, Aliza Zaidman, Amir Nutman, Tahel Ilan Ber, Alon Angel, Lior Kellerman, Eran Barash, Roy Navon, Olga Boico, Yael Israeli, Michal Rosenberg, Amir Gelman, Roy Kalfon, Einav Simon, Noa Avni, Mary Hainrichson, Oren Zarchin, Tanya M. Gottlieb, Kfir Oved, Eran Eden, Boaz Tadmor

## Abstract

The challenge of treating severely ill COVID-19 patients is particularly great due to the need to simultaneously manage oxygenation and the inflammatory state without compromising viral clearance. Currently, there are many tools to aid in oxygen management and in monitoring viral replication. However, predictive biomarkers for monitoring the host immune response across COVID-19 disease stages and specifically, for titrating immunomodulatory therapy are lacking. We utilized a recently cleared platform (MeMed Key™) that enables rapid and easy serial measurement of IP-10, a host protein implicated in lung injury due to viral-induced hyperinflammation. A dynamic clinical decision support protocol was employed for managing SARS-CoV-2 positive patients admitted to a COVID-19 dedicated medical center run by Clalit Health Services. This is the first protocol to include real-time measurements of IP-10 as a potential aid for regulating inflammation. Overall, 502 serial real-time IP-10 measurements were performed on 52 patients recruited between 7^th^ April 2020 to 10^th^ May 2020, with 12 patients admitted to the intensive care unit (ICU). IP-10 levels correlated with increased COVID-19 severity score and ICU admission. Within the ICU admitted patients, the number of days with IP-10 measurements >1,000 pg/ml was associated with mortality. Upon administration of corticosteroid immunomodulatory therapy, a significant decrease in IP-10 levels was observed. Real-time IP-10 monitoring represents a new tool to aid in management and therapeutic decisions relating to the inflammatory status of COVID-19 patients.

## INTRODUCTION

In the majority of SARS-CoV-2 positive patients, the host mounts a localized immune response sufficient to clear the virus from the lungs, following which the immune response recedes and the patient recovers.^1^ In less than 15% of patients, a dysregulated immune response ensues, triggering multiple hyperinflammatory pathways involving a plethora of immune, endothelial, and other cell types.^2,3^ The hyperinflammation involves a release of cytokines and chemokines creating a cytokine release syndrome, which has been implicated in acute lung injury, acute respiratory distress syndrome (ARDS), multiple organ failure and mortality.^4–6^ The cytokine release syndrome resembles secondary hemophagocytic lymphohistocytosis and is characterized by increased interleukin (IL)-6, IL-2, IL-7, granulocyte colony stimulating factor, interferon-γ induced protein 10 (IP-10, also known as CXCL-10), monocyte chemoattractant protein 1, macrophage inflammatory protein 1-α, and tumor necrosis factor-α.^5,7^ Accumulating evidence points specifically to IP-10 as a marker for COVID-19 disease progression, with maintained high levels associated with mortality.^8–13^ In various viral infection models, IP-10 is elevated^14–17^ and implicated initially as promoting viral clearance,^18^ but also, as an effector of immune-mediated acute lung injury, suggesting it plays a key role in both the normal and dysregulated responses to SARS-CoV-2 infection.^19–21^ Accordingly, it has been proposed that IP-10 may represent a marker for COVID-19 disease progression as well as a therapeutic target for preventing lung injury.

The challenge of treating severely ill COVID-19 patients is particularly great due to the need to simultaneously manage oxygenation and the inflammatory state without compromising viral clearance (Figure 1). Accordingly, although accumulating evidence supports suppressing the immune system in severe COVID-19 patients in order to mitigate the cytokine release syndrome,^22^ there is also a concern that over-suppression may reduce the immune system’s capacity to respond effectively to the virus and increase susceptibility to secondary infections.^23^ Corticosteroids in general and methylprednisolone in particular are strong non-specific immunomodulatory agents and their use in severe COVID-19 patients is the topic of ongoing debate and clinical research.^22,23^ Currently, several tools are available for guiding therapeutic decisions on oxygenation and viral replication, but actionable biomarkers for monitoring the host-pathogen interaction and ensuing inflammatory responses are lacking. Clinical monitoring of IP-10 has been restricted in the absence of a rapid diagnostic test. MeMed Key™ is a novel platform recently cleared to provide IP-10 measurements in 15 minutes.

**Figure 1:**
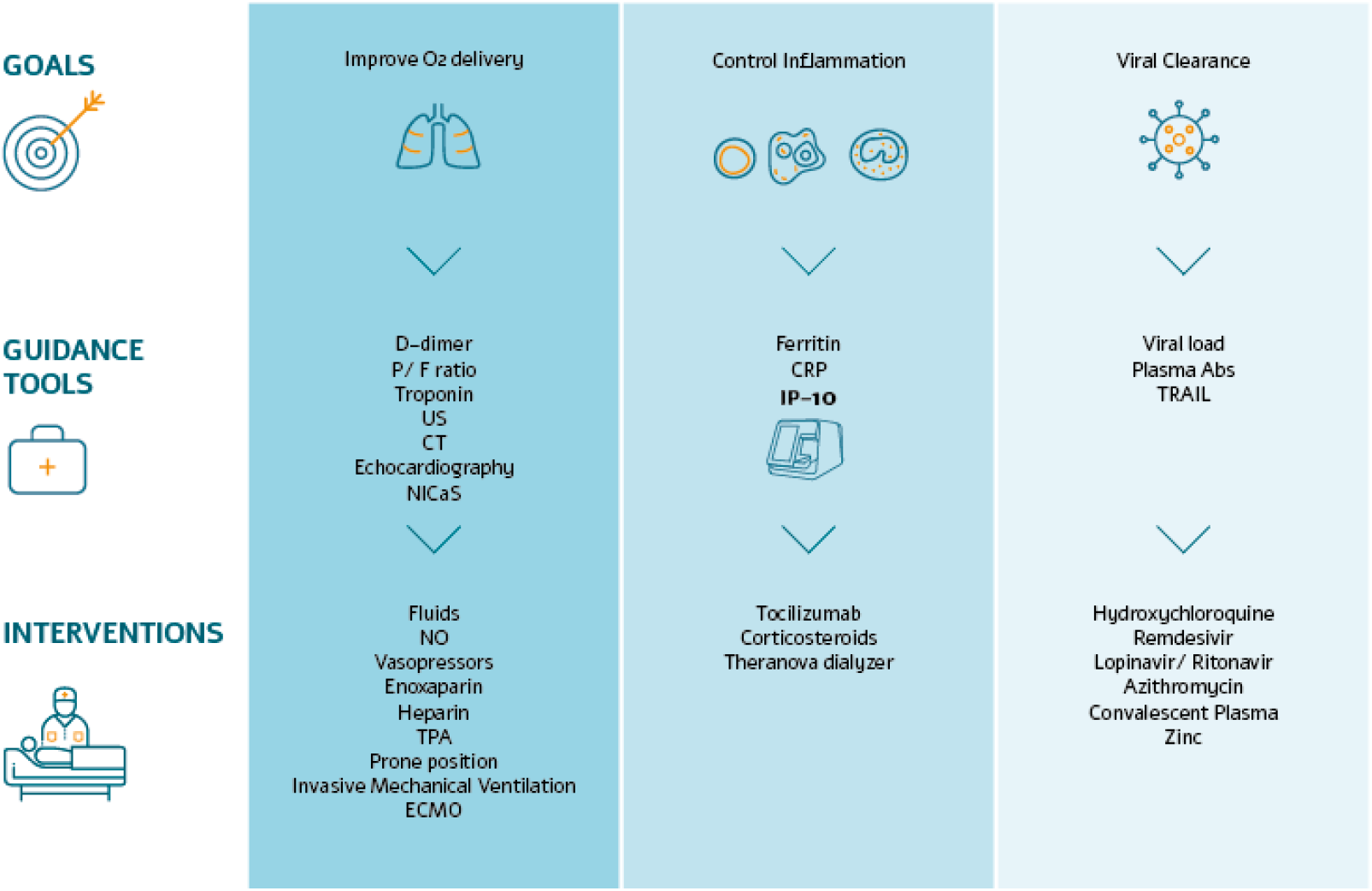
Overview of dynamic clinical decision support protocol employed for managing SARS-CoV-2 patients admitted to a COVID-19 dedicated medical center (Rabin Medical Center, HaSharon, Israel). P/F ratio, Ratio of arterial oxygen partial pressure (PaO2) to fractional inspired oxygen (FiO2); US, Ultrasound; CT, Computed tomography; NICaS, Non-Invasive Cardiac system; NO, Nitric oxide; TPA, Tissue plasminogen activator; ECMO, Extracorporeal membrane oxygenation; CRP, C-reactive protein; IP-10, interferon-γ induced protein 10 (also known as CXCL-10); Plasm Abs, Plasm antibodies; TRAIL, TNF-related apoptosis inducing ligand.

We developed a clinical decision support protocol aimed at achieving 3 predefined therapeutic goals: improving oxygen delivery, controlling inflammation, and promoting viral clearance. Based on our hypothesis that real-time IP-10 measurements can support detection and continuous monitoring of patients with a dysregulated immune response and potentially allow personalized corticosteroid regimens to improve patient outcome, we included longitudinal observation of IP-10 levels in the protocol as a potential tool to aid in inflammation control.

## METHODS

### Study Design

The prospective study included a cohort of SARS-CoV-2 positive patients recruited at a COVID-19 dedicated medical center (n = 52). An additional retrospective study was conducted to enable comparison of IP-10 levels across other acute respiratory infections (n = 182) and healthy subjects (n= 98).

### Patient population

#### COVID-19 patients

The prospective study included 52 SARS-CoV-2 positive patients recruited at a COVID-19 dedicated medical center managed by Clalit Health Services, the second largest Health Management Organization worldwide. All SARS-CoV-2 positive inpatients at the site between 7^th^ April 2020 to 10^th^ May 2020 to whom the clinical decision support protocol was applied were recruited to the study. IRB approval was obtained and informed consent was waived as biomarker measurements were performed on serum remnants from routine blood draws. NCT number: NCT04389645.ARDS and its severity were defined according to the Berlin definition.^24^

#### Patients with other respiratory viruses

A retrospective analysis was performed on a cohort of adult patients (n = 182) pooled from 2 studies, Curiosity^14^ (NCT01917461) and Observer (NCT03011515) who fulfilled all of the following inclusion criteria: meet inclusion/exclusion criteria of original study; detection of at least one of the following viruses (human rhinovirus [HRV], respiratory syncytial virus [RSV], Influenza A [Flu A], Influenza B [Flu B], human coronavirus [HCoV]); and adjudication as viral infection by an expert panel (no bacterial detection).

#### Healthy subjects

A retrospective analysis was performed on a cohort of adult patients (n = 98) from Curiosity^14^ (NCT01917461) and Observer (NCT03011515) who were afebrile with no apparent infectious disease.

### Clinical decision support protocol

An overview of the clinical decision support protocol is provided in Figure 1. In brief, to manage oxygenation, PaO2/FiO2 (P/F, ratio of arterial oxygen partial pressure to fractional inspired oxygen) was monitored twice daily and positive end-expiratory pressure (PEEP) titration was made based on cardiac output measurements when indicated. All patients received high dose N-acetylcysteine and bromohexin inhalations in order to induce mucolysis and prevent sticky secretion and atelectasis. Intra-tracheal injected surfactant was an option as a compassionate therapy for refractory hypoxemic ventilated patients.

Echocardiography and bedside Doppler ultrasound were ordered when pulmonary emboli or deep vein thrombosis were clinically suspected and when D-dimer levels significantly increased. Troponin levels were measured daily and electrocardiogram measurements were performed routinely to detect QT prolongation.

All ICU patients received 40 mg twice daily of enoxaparin. D-dimers were measured daily and the enoxaparin dose was adjusted up to 60 mg twice daily when D-dimer levels increased significantly. In case of proven thrombosis (as visualized by imaging), full anti-coagulation therapy was recommended. When minor bleeding occurred, sub-cutaneous heparin was administered. All ICU patients considered as hyperinflammatory were administered a total tocilizumab dosing of 800 mg, given in 2 separate doses of 400 mg, 12 hours apart. The following parameters were considered as reflective of a hyperinflammatory state: C-reactive protein (CRP) > 90 mg/l, ferritin >500 ng/dl, and IP-10 >1,000 pg/ml. All hyperinflammatory ICU patients were administered methylprednisolone or equivalent dosing of another corticosteroid; basic dosing was 1.5 mg/kg divided into 3 doses. Patients on hemodialysis who were hyperinflamed had access to cytokine removal hemodialysis (Theranova dialyzer).

The anti-viral therapy was based on combinations of hydroxychloroquine, azithromycin, zinc, and lopinavir/ritonavir at the discretion of the infectious disease consultant and was discussed daily.Patients who did not show clinical improvement were given convalescent plasma divided into two doses as a compassionate therapy.

Compassionate Remdesivir became available at the end of April 2020. It was given to only one patient for a total of 7 days (200 mg loading dose followed by 100 mg maintenance dose from day 2 onwards) and was stopped as a result of elevated liver function tests.

### Laboratory procedures

Blood was collected serially from 52 patients as part of routine blood draw into serum separating tubes (SST). Serum was separated within 2 hours of collection and the fresh serum samples were measured on the MeMed Key™ (MeMed, Israel) rapid immunoassay platform that provides measurements of 3 immune system proteins (IP-10, CRP, and TNF-related apoptosis inducing ligand [TRAIL]), in 15 minutes, based on MeMed BV™ (MeMed, Israel) testing cartridges. Any unused serum was frozen at −20°C for up to one week and then transferred for storage at −80°C.

Frozen serum was used to determine retrospectively blood concentrations of IL-6. Measurements were performed using ‘Quantikine human IL-6’ (R&D systems, MN, USA) ELISA assay kits. Due to high IL-6 concentrations, each serum sample was diluted 1:4 and 1:40 with RD6F calibrator diluent supplied in the kits. Results were averaged when the difference in concentration between the 2 dilutions was <20%, otherwise the dilution in the linear range of the calibration curve was taken.Measurements were performed using an Absorbance Microplate Reader Sunrise and Magellan program (Tecan life science Grödig, Austria).

SARS-CoV-2 positivity was determined by RT-PCR (Seegene Inc, South Korea).

### Statistical analysis

Continuous variables are reported as median (interquartile range). Values for 2 related variables were compared using Mann-Whitney-U test (for continuous values) and Fisher’s Exact test (for discrete values). Steroid dosage was converted to methylprednisolone equivalent dosage, using MedCalc’s Steroid Conversion Calculator.^25^ The analysis was performed using Python 3.7.6.

## RESULTS

### Patient population

Overall, 502 serial blood draws were sampled from 52 SARS-CoV-2 patients that were admitted to the COVID-19 dedicated medical center between 7^th^ April 2020 and 10^th^ May 2020 and included in the cohort study, with 12 admitted to the ICU. The median age of the 52 patients was 69, 69% were male, 23% were ventilated, 4 died. There was low in-ICU mortality (1 death, 8%) and non-pulmonary organ failure (0%); the 90-day mortality for ICU patients was 25% (3 deaths). The most common comorbidities were diabetes (42%) and hypertension (46%) (Table 1).

**Table 1:** Demographics of COVID-19 patients (total n = 52), non-ICU (n = 40) and ICU (n = 12)

|  |  | All (n=52) | Non ICU (n=40) | ICU (n=12) | p-value |
| --- | --- | --- | --- | --- | --- |
| General | Age | 68.6 (16.7) | 66.4 (20.0) | 70.2 (13.2) | 0.313 |
|  | Female | 16 (31.0%) | 15 (38.0%) | 1 (8.0%) | 0.078 |
| Smoking Status | Smoker | 3 (6.0%) | 3 (8.0%) | 0 (0.0%) | 1 |
| Other | Death | 4 (8.0%) | 1 (2.0%) | 3 (25.0%) | 0.034 |
|  | Discharged | 51 (98.0%) | 39 (98.0%) | 12 (100.0%) | 1 |
|  | Highest Temp | 37.4 (1.2) | 37.3 (0.9) | 38.3 (0.9) | <0.001 |
|  | Duration of Hospitalization | 11.0 (13.5) | 9.0 (7.5) | 26.5 (12.2) | <0.001 |
|  | Ventilated | 12 (23.0%) | 2 (5.0%) | 10 (83.0%) | <0.001 |
|  | Time Ventilated | 0.0 (0.0) | 0.0 (0.0) | 15.0 (11.0) | <0.001 |
| Co-infection | Bacterial Infection | 10 (19.0%) | 2 (5.0%) | 8 (67.0%) | <0.001 |
|  | Fungal Infection | 4 (8.0%) | 0 (0.0%) | 4 (33.0%) | 0.002 |
| Conditions | Diabetes | 22 (42.0%) | 16 (40.0%) | 6 (50.0%) | 0.74 |
|  | Cardiovascular Disease | 9 (17.0%) | 7 (18.0%) | 2 (17.0%) | 1 |
|  | Cerebrovascular Disease | 3 (6.0%) | 3 (8.0%) | 0 (0.0%) | 1 |
|  | Dyslipidemia | 9 (17.0%) | 6 (15.0%) | 3 (25.0%) | 0.415 |
|  | Hypertension | 24 (46.0%) | 18 (45.0%) | 6 (50.0%) | 1 |
|  | Hypothyroidism | 6 (12.0%) | 2 (5.0%) | 4 (33.0%) | 0.021 |
|  | Malignancy | 5 (10.0%) | 2 (5.0%) | 3 (25.0%) | 0.074 |
|  | Obesity (BMI>30 kg/m2) | 9 (17.0%) | 8 (20.0%) | 1 (8.0%) | 0.666 |
| Symptoms | Sore Throat | 1 (2.0%) | 1 (2.0%) | 0 (0.0%) | 1 |
|  | Sputum Production | 1 (2.0%) | 1 (2.0%) | 0 (0.0%) | 1 |
|  | Weakness | 12 (23.0%) | 11 (28.0%) | 1 (8.0%) | 0.253 |
|  | Rhinorrhea | 1 (2.0%) | 1 (2.0%) | 0 (0.0%) | 1 |
|  | Chest pain | 2 (4.0%) | 2 (5.0%) | 0 (0.0%) | 1 |
|  | Diarrhea | 6 (12.0%) | 6 (15.0%) | 0 (0.0%) | 0.316 |
|  | Dyspnea | 23 (44.0%) | 17 (42.0%) | 6 (50.0%) | 0.746 |
|  | Fever | 29 (56.0%) | 18 (45.0%) | 11 (92.0%) | 0.007 |
|  | Loss of Taste and Smell | 5 (10.0%) | 5 (12.0%) | 0 (0.0%) | 0.578 |
|  | Cough | 26 (50.0%) | 17 (42.0%) | 9 (75.0%) | 0.097 |
| Markers | AST (GOT) (u/l) (max) | 37.0 (38.8) | 28.4 (25.5) | 86.0 (91.2) | <0.001 |
|  | ALT (GPT) (u/l) (max) | 34.2 (70.1) | 28.6 (27.0) | 120.1 (147.1) | <0.001 |
|  | Glucose (mg/dl) (max) | 153.5 (129.6) | 126.4 (85.1) | 247.0 (76.9) | <0.001 |
|  | LDH (u/l) (max) | 610.0 (347.5) | 500.0 (244.5) | 912.5 (263.8) | <0.001 |
|  | Lymph.abs (K/micl) (min) | 0.8 (0.7) | 0.8 (0.7) | 0.4 (0.4) | 0.003 |
|  | Neu.abs (K/micl) (max) | 5.6 (5.6) | 5.0 (3.0) | 13.8 (6.2) | <0.001 |
|  | PLT (10<sup>3</sup>) (min) | 186.5 (111.8) | 228.0 (123.5) | 139.5 (49.8) | 0.001 |
|  | PaO2/FiO2 (min) | 83.0 (56.5) | 141.0 (0.0) | 80.5 (42.0) | <0.001 |
|  | Troponin (max) | 19.5 (44.5) | 11.5 (18.5) | 69.5 (55.5) | <0.001 |
|  | Urea (max) | 46.3 (40.6) | 37.2 (27.5) | 105.0 (66.9) | <0.001 |
|  | Ferritin (max) | 595.0 (1022.8) | 462.0 (459.2) | 1691.5 (916.8) | <0.001 |
|  | D-Dimer (max) | 1319.0 (4050.0) | 1140.0 (1377.5) | 7235.5 (18598.2) | <0.001 |
|  | Creatinine (mg/dl) (max) | 1.0 (0.4) | 0.9 (0.4) | 1.0 (0.5) | 0.086 |
|  | Total Bilirubin (mg/dl) (max) | 0.5 (0.5) | 0.5 (0.4) | 0.9 (0.5) | 0.001 |
|  | Total Protein (g/dl) (max) | 7.0 (0.7) | 7.0 (0.6) | 7.0 (0.8) | 0.136 |
|  | CRP (max) | 97.9 (122.8) | 95.0 (92.1) | 158.4 (172.6) | 0.04 |
| Scores | COVID-19 severity | 26 (50.0%) | 14 (35.0%) | 12 (100.0%) | 0 |
|  | qSOFA (on admission) | 0.0 (1.0) | 0.0 (1.0) | 1.0 (1.2) | 0.003 |
| Treatments | Methylprednisolone | 12 (23.0%) | 1 (2.0%) | 11 (92.0%) | <0.001 |
|  | Hydrocortisone | 10 (19.0%) | 4 (10.0%) | 6 (50.0%) | 0.006 |
|  | Methylprednisolone+Prednisone+Hydrocortisone | 22 (42.0%) | 10 (25.0%) | 12 (100.0%) | 0 |
|  | Renal Replacement Therapy | 2 (4.0%) | 1 (2.0%) | 1 (8.0%) | 0.412 |
|  | Remdesivir | 1 (2.0%) | 0 (0.0%) | 1 (8.0%) | 0.231 |
|  | Prednisone | 6 (12.0%) | 6 (15.0%) | 0 (0.0%) | 0.316 |
|  | Nitric Oxide | 5 (10.0%) | 0 (0.0%) | 5 (42.0%) | <0.001 |
|  | ECMO | 1 (2.0%) | 0 (0.0%) | 1 (8.0%) | 0.231 |
|  | Vasopressors | 11 (21.0%) | 2 (5.0%) | 9 (75.0%) | <0.001 |
|  | Convalescent Plasma | 7 (13.0%) | 3 (8.0%) | 4 (33.0%) | 0.041 |
|  | Azithromycin | 35 (67.0%) | 24 (60.0%) | 11 (92.0%) | 0.076 |
|  | Antiviral Lopinavir/Ritonavir | 7 (13.0%) | 1 (2.0%) | 6 (50.0%) | <0.001 |
|  | Antiviral Hydroxychloroquine plus Azithromycin | 33 (63.0%) | 22 (55.0%) | 11 (92.0%) | 0.037 |
|  | Antiviral Hydroxychloroquine | 36 (69.0%) | 24 (60.0%) | 12 (100.0%) | 0.01 |
|  | Antibiotics | 30 (58.0%) | 19 (48.0%) | 11 (92.0%) | 0.008 |
|  | Tocilizumab | 13 (25.0%) | 3 (8.0%) | 10 (83.0%) | <0.001 |
|  | Days Under Systemic Corticosteroids | 0.0 (6.2) | 0.0 (0.2) | 7.5 (7.5) | <0.001 |
| First BV Test | TRAIL | 56.6 (35.4) | 64.0 (27.4) | 33.8 (12.9) | 0.001 |
|  | CRP | 70.9 (85.4) | 77.8 (70.1) | 36.0 (166.8) | 0.5 |
|  | IP-10 | 596.3 (1213.1) | 502.3 (895.8) | 1642.2 (2155.8) | 0.019 |

Among the 12 ICU patients, 10 needed invasive mechanical ventilation and exhibited a pulmonary status compatible with severe ARDS (Table 2). Of these, 7 were weaned during their ICU stay and transferred to a rehabilitation center, 1 was transferred to a respiratory rehabilitation center (Patient #5) and 2 (Patients #3 and#9) were transferred to the internal medicine department, where patient #3 died. 8 patients developed secondary bacterial infection (Table 2).

**Table 2:**
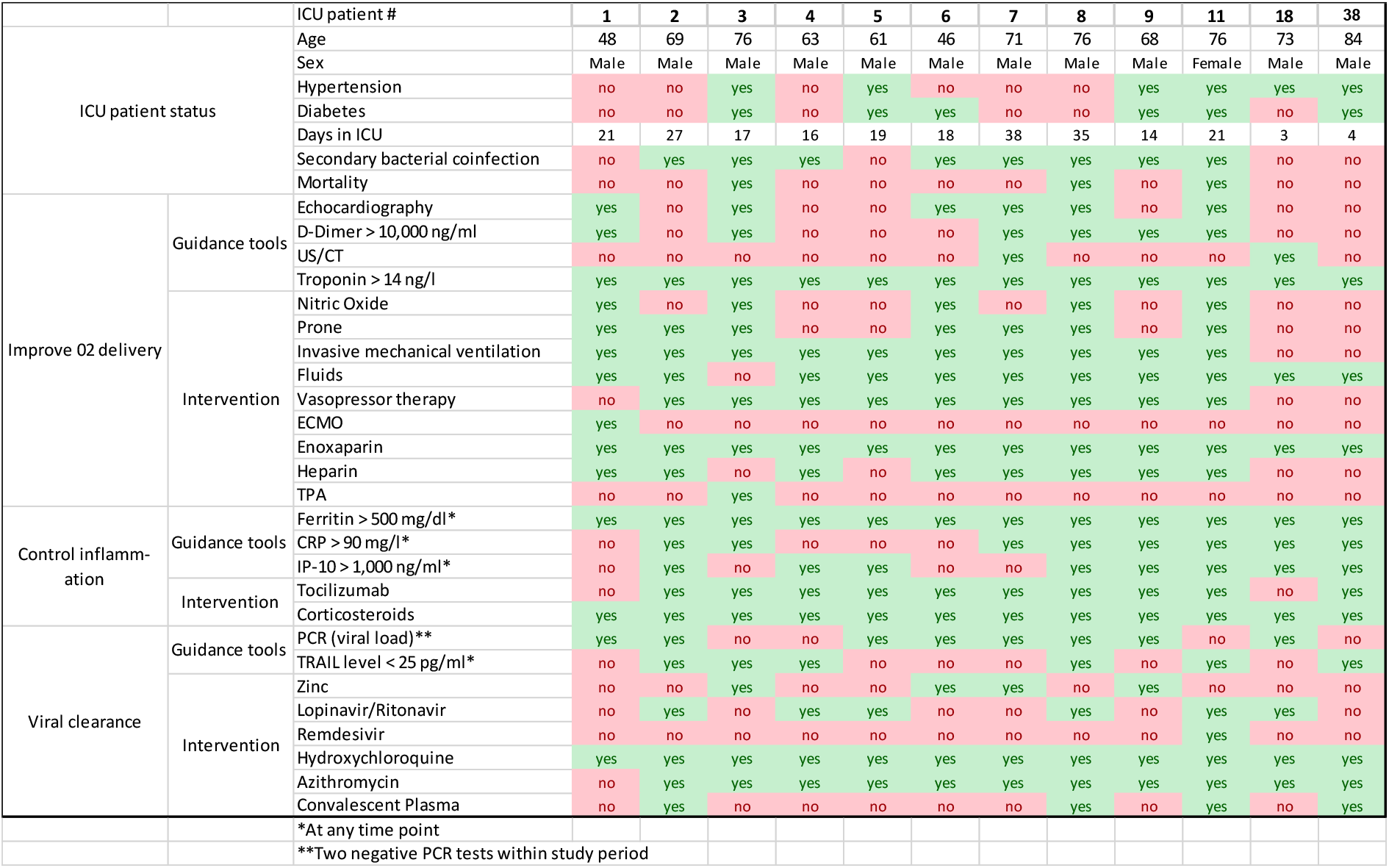
Status of the 12 ICU patients

### IP-10 levels indicate hyperinflammation and associate with mortality

A dynamic clinical decision support protocol that includes IP-10 was employed for managing the SARS-CoV-2 patients admitted to the COVID-19 dedicated medical center (Figure 1). Based on an evolving understanding of the complex host-pathogen interaction during disease progression, the goals of this protocol were to improve oxygen delivery and control inflammation without compromising viral clearance. Observations relating to improving oxygen delivery are described in the Supplementary material.

To assess IP-10 levels in SARS-CoV-2 infection as compared to other viral infections and healthy subjects, a retrospective analysis was performed that included cohorts from previous studies (Figure 2). IP-10 levels are increased in patients with viral infections versus healthy subjects (p<0.001). Among the viral infections, IP-10 levels are higher when the infection is associated with pulmonary pathology (RSV, Flu A, Flu B, HCoV, and SARS-Cov-2), versus non-pulmonary pathology (HRV; p < 0.02). Moreover, among the SARS-CoV-2 positive patients, IP-10 is elevated greatly in ICU versus non-ICU patients (p<0.019).

**Figure 2:**
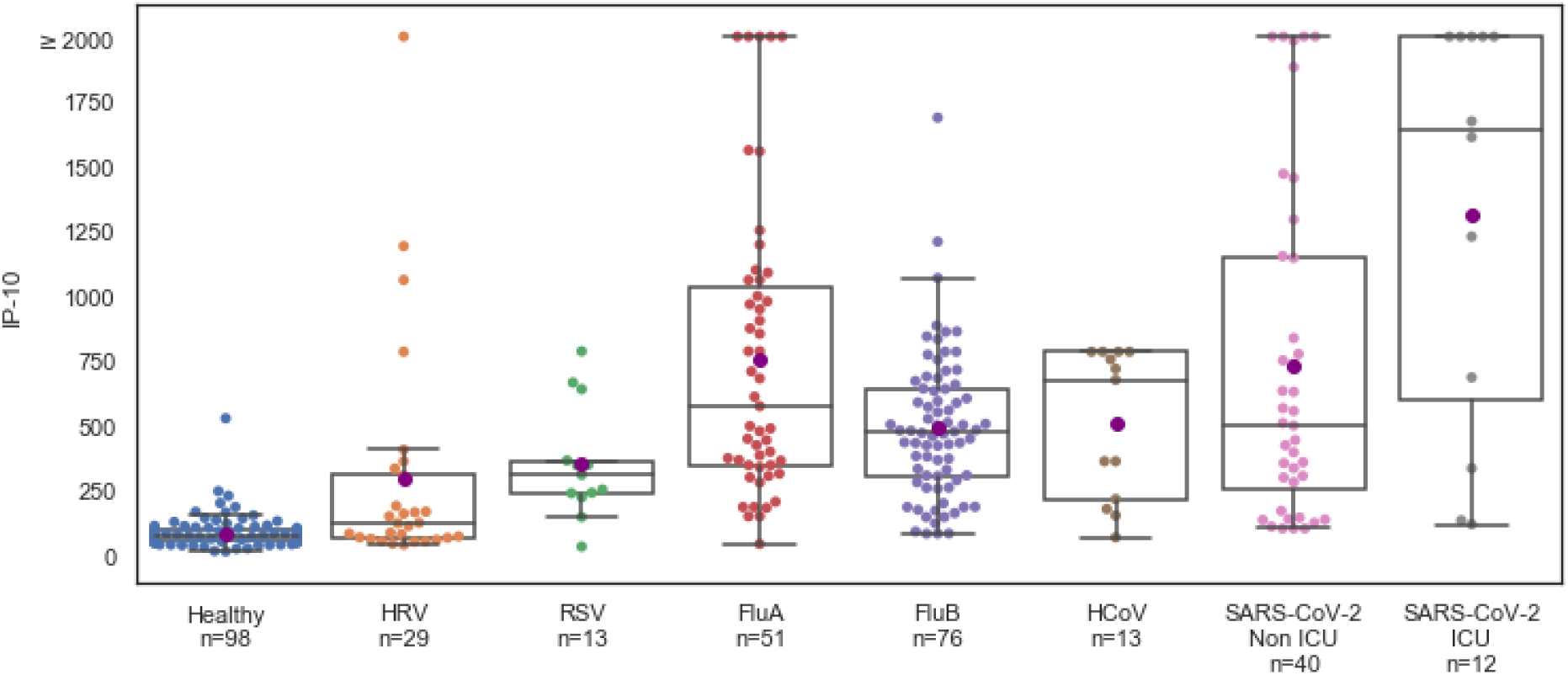
IP-10 levels are significantly higher in viral infections associated with pulmonary pathology. IP-10 levels in the non-ICU (n = 40) and ICU (n = 12) COVID-19 patients and IP-10 levels in healthy subjects (n = 98) and virally infected patients recruited in Curiosity^14^ (NCT01917461) and Observer (NCT03011515) studies (n = 182) with PCR-confirmed viral infections including human rhinovirus (HRV), respiratory syncytial virus (RSV), influenza (Flu), and human coronavirus (HCoV). Each circle represents a patient. Black line corresponds to the group median and purple circle corresponds to group average. The box indicates patients with values between the 25 and 75 percentiles. The whiskers indicate patients with values between 2.5 and 97.5 percentiles. For the COVID-19 patients, IP-10 was measured using MeMed Key™ at multiple time points and the first IP-10 measurement is shown.

Focusing on the prospective cohort of 52 SARS-CoV-2 positive patients, IP-10 >1,000 pg/ml correlated with increased COVID-19 severity score^5^ (p<0.01) and ICU admission (p<0.05; Supplementary Table 1). Among the 12 ICU patients, 11 were considered hyperinflammatory, and 10 received tocilizumab and methylprednisolone; patient #18 was admitted to the ICU after pneumothorax surgery and was not administered tocilizumab (Table 2). For COVID-19 patients (n = 52) and the sub-group of ICU patients (n = 12), the number of days with IP-10 > 1,000 pg/ml was associated with mortality (Figure 3). In contrast, the number of days with CRP > 90 mg/l or ferritin > 500 mg/dl was not associated with mortality (Supplementary Figure 1). Overall, 3 ICU patients died; patient #3 died of candidemia. Focusing on the remaining 2 ICU patients who died, IP-10 > 1,000 pg/ml was exhibited for an average of 7.5 days (standard deviation [SD], 2.1 days) in contrast to the 9 surviving patients who exhibited IP-10 > 1,000 pg/ml for an average of 1 day (SD, 1.1 day; p=0.04).

**Figure 3:**
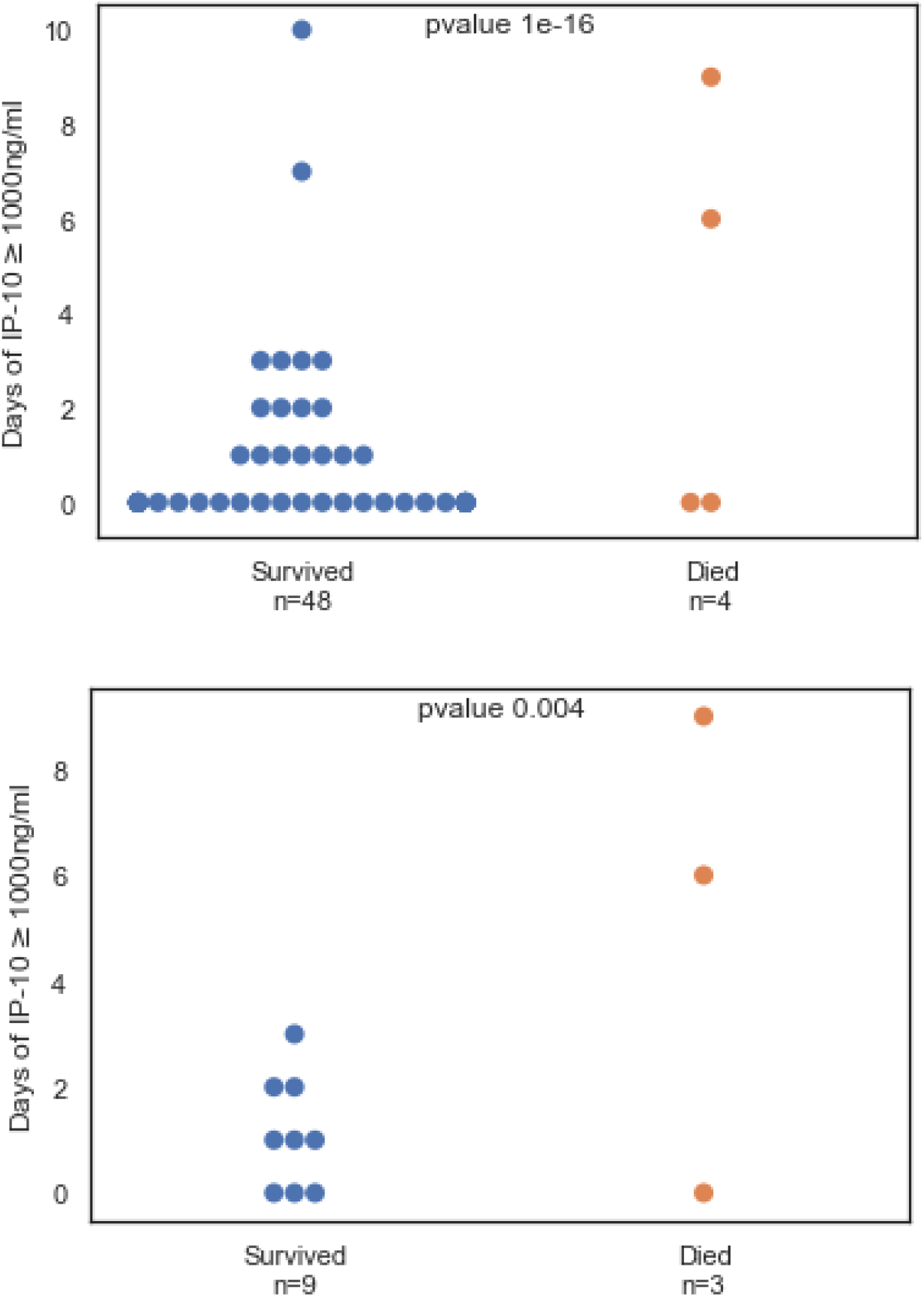
Mortality correlates with the number of days IP-10 >1000 pg/ml in SARS-CoV-2 positive patients (n = 52, upper panel) and among the subset of ICU patients (n = 12, lower panel). IP-10 was measured using MeMed Key™ at multiple time points during the SARS-CoV-2 positive patient’s hospitalization. At least one IP-10 measurement ≥ 1,000 pg/ml in a given day was sufficient to classify as a day > 1,000 pg/ml. Each circle represents a patient; blue dots represent patients who survived and red dots represent patients who died. The cause of death of the ICU patient with a low number of days of IP-10 > 1,000 pg/ml was candidemia and of the non-ICU patient was metastatic breast cancer.

All ICU patients exhibited reduced IP-10 levels within 3 days of their first corticosteroid treatment administered in the ICU (Figure 4). In the case where patients were given additional corticosteroids a decrease in IP-10 levels was observed (Figure 5); patients #8 and #11 who exhibited multiple days of highly elevated IP-10 died. Retrospective examination of IL-6 levels in the ICU patients revealed a weak correlation with IP-10 levels (Pearson correlation coefficient, 0.26) and no association with mortality, both for number of days with IL-6 > 80 pg /ml or IL-6 > 100 pg/ml (Supplementary Figure 2).

**Figure 4:**
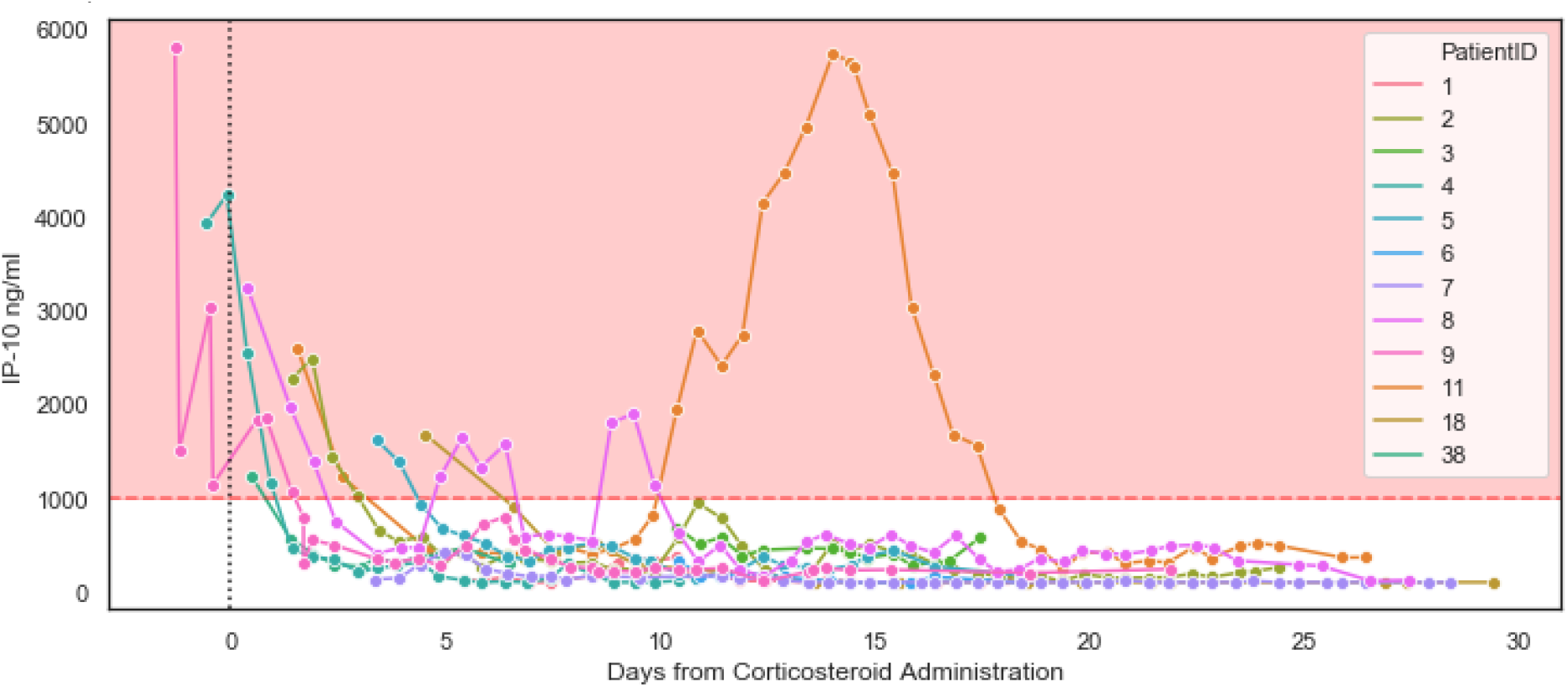
Initiation of corticosteroid therapy was reflected by a decrease in IP-10 levels (ICU patients, n = 12) IP-10 was measured using MeMed Key™ at the indicated time points during the stay of the patients in the ICU. Day 0 = initiation of corticosteroid therapy. Pink area indicates IP-10 ≥ 1,000 pg/ml. Patients #2, #8 and #11 exhibited subsequent surges in IP-10 expression and are detailed in Figure 5.

**Figure 5:**
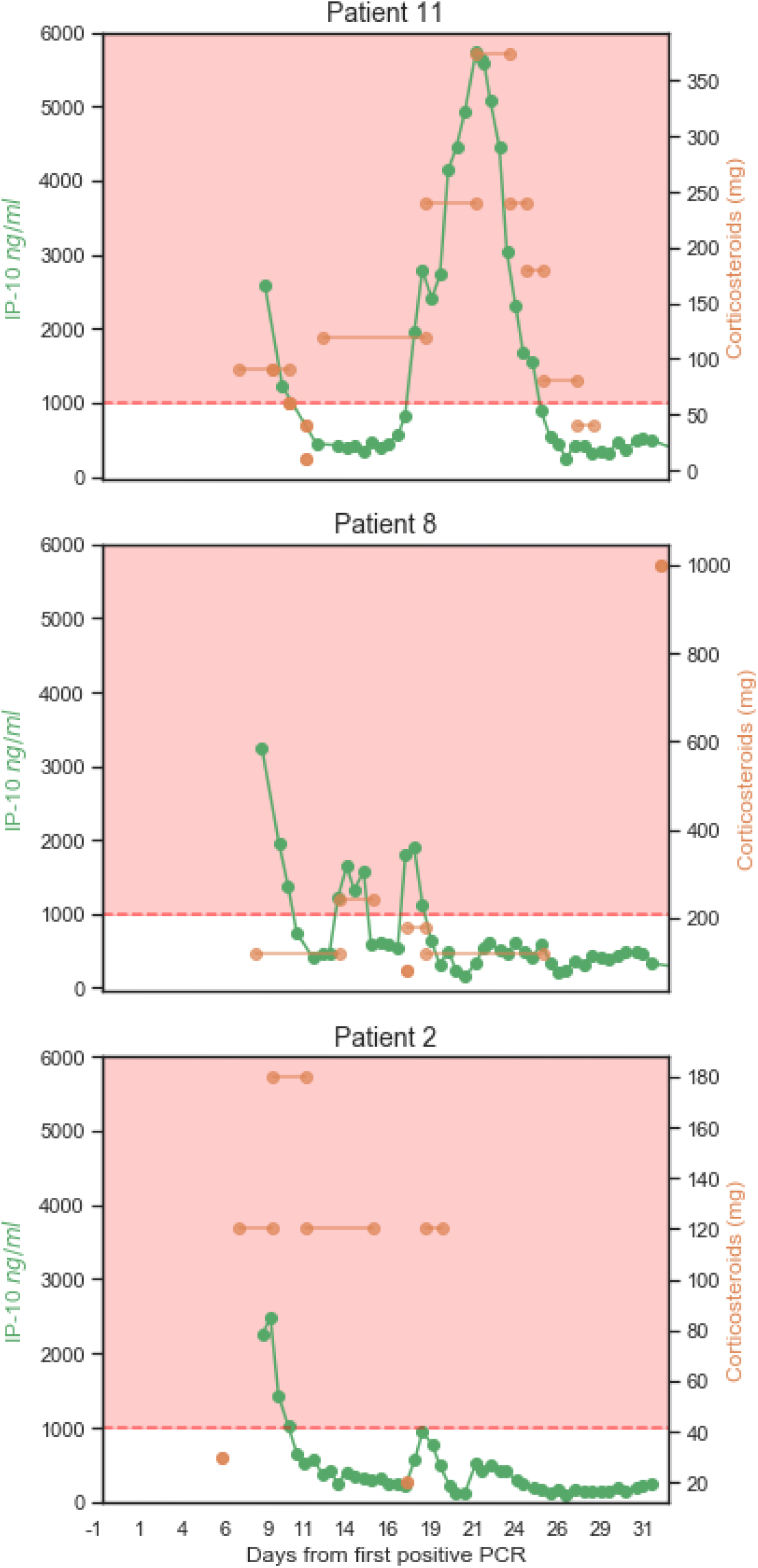
IP-10 levels reflect personalized corticosteroid dosing (both increase and decrease) These 3 patients had a relapse of IP-10 levels >1,000 pg/ml, after initial treatment with corticosteroids. Right Y-axis shows the normalized^25^ levels of corticosteroids administered (hydrocortisone was converted to methylprednisolone). Left Y-axis shows the levels of IP-10 measured by MeMed Key™ at the indicated time points. X-axis shows the days from the first positive SARS-CoV-2 PCR. Lines connecting the doses indicate that the dose was given in each of the intervening day. Pink area indicates IP-10 > 1,000 pg/ml. Patient 2 survived, patients 8 and 11 died.

### TRAIL levels flag viral persistence

The MeMed Key™ platform is also cleared to provide measurements of the immune protein TRAIL. Applying the formal criterion of two negative SARS-CoV-2 PCR tests as confirmation of viral clearance, eight out of the 12 ICU patients were clear of the virus within the study period. TRAIL ≤ 25 pg/ml was associated with longer duration of SARS-CoV-2 positivity (p = 0.012). The number of days TRAIL ≤ 25 pg/ml was found to correlate with the number of virus positive days (Pearson correlation coefficient, 0.7).

## DISCUSSION

This is the first study to examine the potential utility of rapid and serial IP-10 measurements as a tool within a clinical decision support protocol aimed at improving oxygen delivery and controlling inflammation without increasing susceptibility to secondary infections or compromising viral clearance. The study was prompted by the need for additional, specific tools to monitor and assess COVID-19 patient inflammatory status. There is mounting evidence that aberrantly high IP-10 levels play a role in the acute lung injury that is observed in COVID-19 patients and that abnormally elevated IP-10 contributes to mortality. In line with this emerging understanding of the participation of IP-10 in the complex and multi-factorial COVID-19 pathophysiology, we found that IP-10 levels are associated with disease severity and reflect the patient’s response to corticosteroids. Specifically, we observed that relapsing surges of abnormally elevated IP-10 expression were characteristic of the patients that subsequently died, possibly due to more extensive lung damage. These findings suggest that longitudinal and real-time IP-10 measurements could help with personalizing immunomodulatory treatment regimens for COVID-19 patients and may support better patient outcomes.

Our clinical decision support protocol was developed in parallel to the Yale New Haven YNHHS treatment algorithm (latest version June 22)^26^ and the Massachusetts General Hospital COVID-19 treatment guidance (June 24).^27^ Similar to our protocol, the latest YNHHS algorithm recommends interventions to address oxygenation, inflammation, and viral clearance. Notably, in the YNHHS algorithm for hospitalized adult patients with severe disease, it is recommended that a cytokine panel be conducted on ICU admission and steroids be given at the discretion of the primary team. We propose that the information provided by multiple IP-10 measurements performed using an easy and rapid measurement platform, as opposed to a one-time cytokine panel, serves as actionable and dynamic support to steroid treatment decision-making by the primary team.

Currently, the non-specific inflammatory biomarkers CRP and ferritin are employed to detect and assess the inflammatory status of COVID-19 patients. IP-10 represents a complementary tool that has the added advantage of rapidly indicating response to even a single dose of corticosteroid therapy, enabling its use as an aid to personalize corticosteroid treatment regimens. In addition, unlike CRP and ferritin, IP-10 exhibited a strong correlation with mortality for the ICU patients, likely reflecting its purported role as an effector of acute lung injury in COVID-19 disease progression. Of note, this study did not include many COVID-19 patients at early stages of disease progression. We hypothesize that serial measurement of IP-10 levels in such patients may flag those who are transitioning to the hyperinflamed state and enable early initiation of corticosteroids in this subset of SARS-CoV-2 positive patients. Further studies are warranted to address this premise directly. Another marker used to monitor inflammation is IL-6, which is the therapeutic target of tocilizumab. Its use as an indicator of inflammatory status may be constrained when the patients are under tocilizumab therapy.^28^ In line with previous reports, we did not find that IL-6 expression is an indicator of mortality.^23^

A recent meta-analysis including 24 observational studies from across the world (Asia, Europe, and North America) involving 10,150 COVID-19 patients demonstrated a combined in-ICU mortality of 41.6% (95% confidence interval [CI], 34.0-49.7%). We did not observe non-pulmonary organ failure and the in-ICU mortality rate was 8%. The main limitation of our study is that the study design was not a randomized controlled trial, which limits the ability to estimate the magnitude of improvement in patient outcome that is attributable to implementation of the dynamic clinical decision support protocol. Moreover, the cohort study design does not allow evaluation of the contribution of different elements of the protocol, in particular, the personalized corticosteroid regimens. In addition, this is a single-center study; application of the protocol at additional sites and across multiple cohorts is warranted. The pivotal strength of our study is that it is the first prospective investigation of the potential value of serial and rapid IP-10 measurements as an indicator of the inflammatory status of COVID-19 patients. In addition, preliminary data was collected suggesting that low TRAIL levels, reported as an indicator of infection severity^29^, when sustained may flag failure to clear the virus. Further studies of TRAIL and its prognostic value are required.

In conclusion, our study supports the utility of using serial and rapid IP-10 measurements as a practical new tool in management of COVID-19 patients. Future studies are warranted to establish the contribution of IP-10 within a clinical decision support protocol to improved COVID-19 patient outcome.

## Data Availability

The data will not be publicly available.

## SUPPLEMENTARY MATERIALS

### Supplementary Results

#### Goal 1: Improve oxygen delivery

Of the 12 ICU patients, nine patients had PEEP above 12 cm/H2O or oliguria or hypotension. Patient #3 displayed low cardiac index (2.2 L/minute) and D-dimers above 30,000 ng/ml and RV dysfunction; this patient was administered a mini dose of tissue plasminogen activator (TPA) but no improvement in oxygenation was observed and the patient died. Patient#2 also displayed low CO measurements (below 2.4 L/minute) and the echocardiography revealed RV dysfunction; this patient did not receive TPA due to mucosal bleeding. A bolus of intra-tracheal surfactant was administered and led to a transient improvement in oxygenation and the patient survived. All ICU patients exhibited elevated D-dimer levels above 1,000 ng/ml. Patient #7 developed colonic perforation with peritonitis and had small vessel thrombosis (in pathology) and a D-dimer level of 76,000 ng/ml.

**Supplementary Table 1:** Demographics of COVID-19 patients (total n = 52), IP-10 < 1,000 pg/ml (n = 33) and IP-10 > 1,000 pg/ml (n = 19) BMI, Body mass index; AST, Aspartate transaminase; AL, Alanine transaminase; LDH, Lactate dehydrogenase; Lymph. Abs, Absolute lymphocytes; Neu. Abs, Absolute neutrophils; PLT, platelets; PaO2/FiO2, Ratio of arterial oxygen partial pressure to fractional inspired oxygen; ECMO, Extracorporeal membrane oxygenation; qSOFA, quick sequential organ failure assessment; TRAIL, TNF-related apoptosis inducing ligand; IP-10, interferon-γ induced protein 10 (also known as CXCL-10); CRP, C-reactive protein; BV Test, serum measurement of TRAIL, CRP and IP-10 performed using MeMed Key™.

|  |  | 0<IP-10<=1000 (n=33) | 1000<IP-10 (n=19) | p-value |
| --- | --- | --- | --- | --- |
| General | Age | 68.8 (21.5) | 68.5 (14.2) | 0.388 |
|  | Female | 12 (36.0%) | 4 (21.0%) | 0.353 |
| Smoking Status | Smoker | 1 (3.0%) | 2 (11.0%) | 0.546 |
| Other | Death | 2 (6.0%) | 2 (11.0%) | 0.617 |
|  | Discharged | 32 (97.0%) | 19 (100.0%) | 1 |
|  | Highest Temp | 37.2 (1.0) | 38.1 (1.3) | 0.001 |
|  | Duration of Hospitalization | 9.0 (13.8) | 12.0 (15.0) | 0.052 |
|  | Ventilated | 6 (18.0%) | 6 (32.0%) | 0.317 |
|  | Time Ventilated | 0.0 (0.0) | 0.0 (12.5) | 0.174 |
| Co-infection | Bacterial Infection | 4 (12.0%) | 6 (32.0%) | 0.142 |
|  | Fungal Infection | 2 (6.0%) | 2 (11.0%) | 0.617 |
| Conditions | Diabetes | 13 (39.0%) | 9 (47.0%) | 0.771 |
|  | Cardiovascular Disease | 7 (21.0%) | 2 (11.0%) | 0.458 |
|  | Cerebrovascular Disease | 3 (9.0%) | 0 (0.0%) | 0.291 |
|  | Dyslipidemia | 4 (12.0%) | 5 (26.0%) | 0.26 |
|  | Hypertension | 11 (33.0%) | 13 (68.0%) | 0.021 |
|  | Hypothyroidism | 2 (6.0%) | 4 (21.0%) | 0.175 |
|  | Malignancy | 1 (3.0%) | 4 (21.0%) | 0.054 |
|  | Obesity (BMI>30 kg/m2) | 7 (21.0%) | 2 (11.0%) | 0.458 |
| Symptoms | Sore Throat | 1 (3.0%) | 0 (0.0%) | 1 |
|  | Sputum Production | 1 (3.0%) | 0 (0.0%) | 1 |
|  | Weakness | 8 (24.0%) | 4 (21.0%) | 1 |
|  | Rhinorrhea | 1 (3.0%) | 0 (0.0%) | 1 |
|  | Chest pain | 2 (6.0%) | 0 (0.0%) | 0.527 |
|  | Diarrhea | 4 (12.0%) | 2 (11.0%) | 1 |
|  | Dyspena | 12 (36.0%) | 11 (58.0%) | 0.157 |
|  | Fever | 16 (48.0%) | 13 (68.0%) | 0.247 |
|  | Loss of Taste and Smell | 4 (12.0%) | 1 (5.0%) | 0.641 |
|  | Cough | 15 (45.0%) | 11 (58.0%) | 0.565 |
| Markers | AST (GOT) (u/l) (max) | 26.6 (35.2) | 55.7 (37.9) | 0.005 |
|  | ALT (GPT) (u/l) (max) | 28.6 (46.1) | 63.4 (85.1) | 0.017 |
|  | Glucose (mg/dl) (max) | 131.2 (110.8) | 184.4 (150.3) | 0.017 |
|  | LDH (u/l) (max) | 491.0 (312.8) | 689.0 (266.0) | 0.011 |
|  | Lymph.abs (K/micl) (min) | 0.9 (0.7) | 0.5 (0.4) | 0.004 |
|  | Neu.abs (K/micl) (max) | 5.1 (3.8) | 8.0 (8.5) | 0.014 |
|  | PLT (10^3) (min) | 229.0 (129.0) | 162.0 (55.0) | 0.013 |
|  | PaO2/FiO2 (min) | 83.0 (63.0) | 89.5 (46.5) | 0.44 |
|  | Troponin (max) | 12.0 (33.0) | 25.0 (40.0) | 0.059 |
|  | Urea (max) | 34.6 (38.0) | 58.0 (57.9) | 0.002 |
|  | Ferritin (max) | 449.4 (697.1) | 901.0 (1147.6) | 0.005 |
|  | D-Dimer (max) | 1173.5 (2575.2) | 2175.0 (6061.5) | 0.055 |
|  | Creatinine (mg/dl) (max) | 0.9 (0.4) | 1.0 (0.4) | 0.008 |
|  | Total Bilirubin (mg/dl) (max) | 0.4 (0.4) | 0.8 (0.6) | 0.018 |
|  | Total Protein (g/dl) (max) | 7.0 (0.7) | 7.0 (0.7) | 0.406 |
|  | CRP (max) | 66.1 (82.3) | 169.5 (109.0) | <0.001 |
| Scores | COVID-19 severity | 11 (33.0%) | 15 (79.0%) | 0.003 |
|  | qSOFA (on admission) | 0.0 (1.0) | 1.0 (1.0) | 0.064 |
| Treatments | Methylprednisolone | 4 (12.0%) | 8 (42.0%) | 0.019 |
|  | Hydrocortisone | 5 (15.0%) | 5 (26.0%) | 0.467 |
|  | Methylprednisolone+Prednisone+Hydrocortisone | 10 (30.0%) | 12 (63.0%) | 0.04 |
|  | Renal Replacement Therapy | 1 (3.0%) | 1 (5.0%) | 1 |
|  | Remdesivir | 0 (0.0%) | 1 (5.0%) | 0.365 |
|  | Prednisone | 3 (9.0%) | 3 (16.0%) | 0.656 |
|  | Nitric Oxide | 2 (6.0%) | 3 (16.0%) | 0.342 |
|  | ECMO | 1 (3.0%) | 0 (0.0%) | 1 |
|  | Vasopressors | 5 (15.0%) | 6 (32.0%) | 0.181 |
|  | Convalescent Plasma | 1 (3.0%) | 6 (32.0%) | 0.007 |
|  | Azithromycin | 16 (48.0%) | 19 (100.0%) | <0.001 |
|  | Antiviral Lopinavir/Ritonavir | 0 (0.0%) | 7 (37.0%) | <0.001 |
|  | Antiviral Hydroxychloroquine plus Azithromycin | 14 (42.0%) | 19 (100.0%) | <0.001 |
|  | Antiviral Hydroxychloroquine | 17 (52.0%) | 19 (100.0%) | <0.001 |
|  | Antibiotics | 15 (45.0%) | 15 (79.0%) | 0.023 |
|  | Tocilizumab | 5 (15.0%) | 8 (42.0%) | 0.047 |
|  | Days Under Systemic Corticosteroids | 0.0 (3.0) | 5.0 (10.5) | 0.007 |
| First BV Test | TRAIL | 58.2 (22.5) | 50.7 (52.7) | 0.131 |
|  | CRP | 38.4 (70.4) | 107.4 (88.8) | <0.001 |
|  | IP-10 | 333.2 (419.5) | 1984.5 (1525.6) | <0.001 |

**Supplementary Figure 1:**
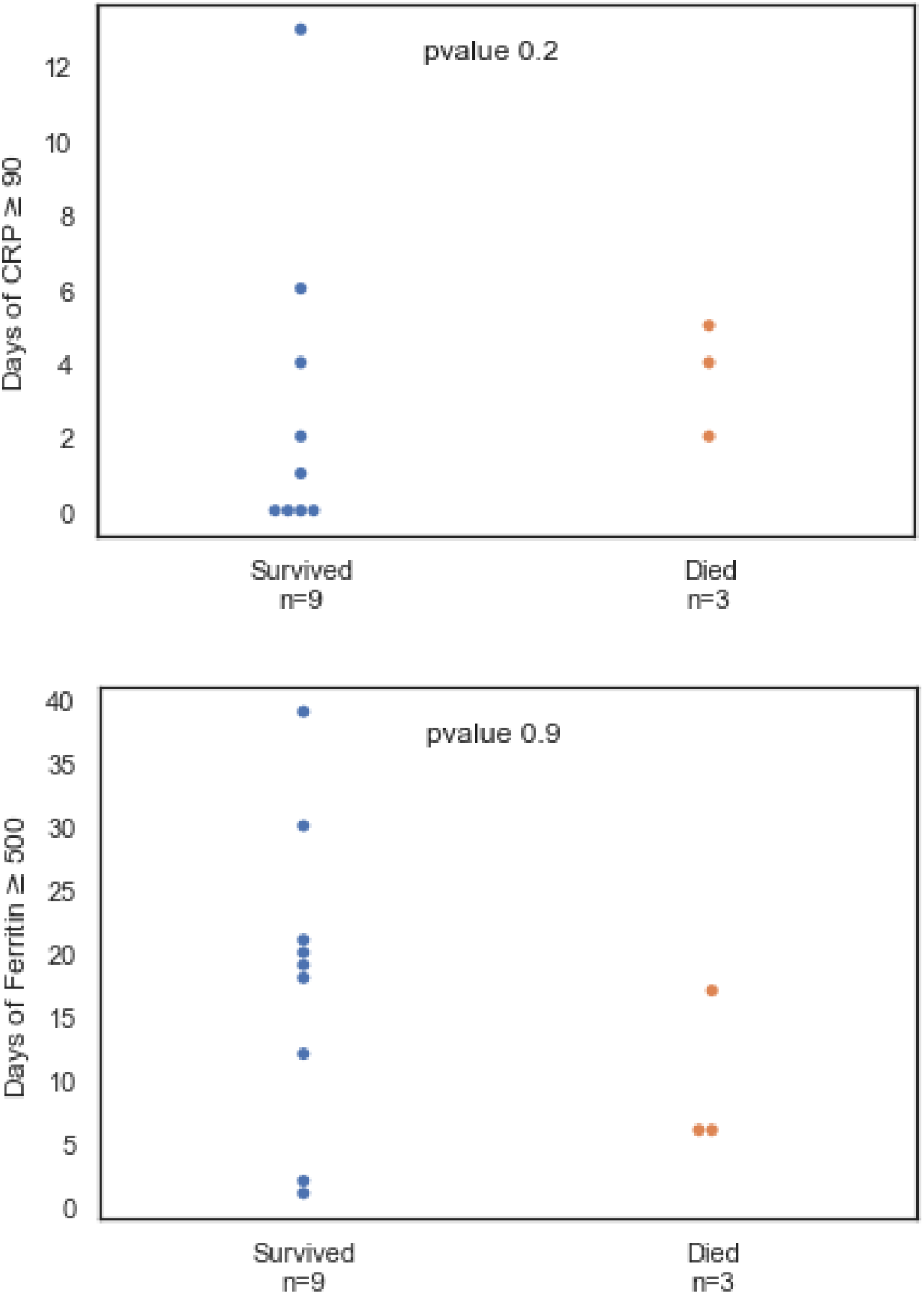
Mortality is not correlated with the number of days CRP > 90 mg/l (upper panel) or number of days ferritin > 500 mg/dl (lower panel) in the ICU patients (n = 12) CRP and ferritin were measured at multiple time points during the 12 patient’s stay in the ICU. At least one CRP > 90 mg/l or > 500 mg/dl in a given day was sufficient for the analysis. Each circle represents a patient; blue dots represent patients who survived (n = 9) and red dots represent patients who died (n = 3).

**Supplementary Figure 2:**
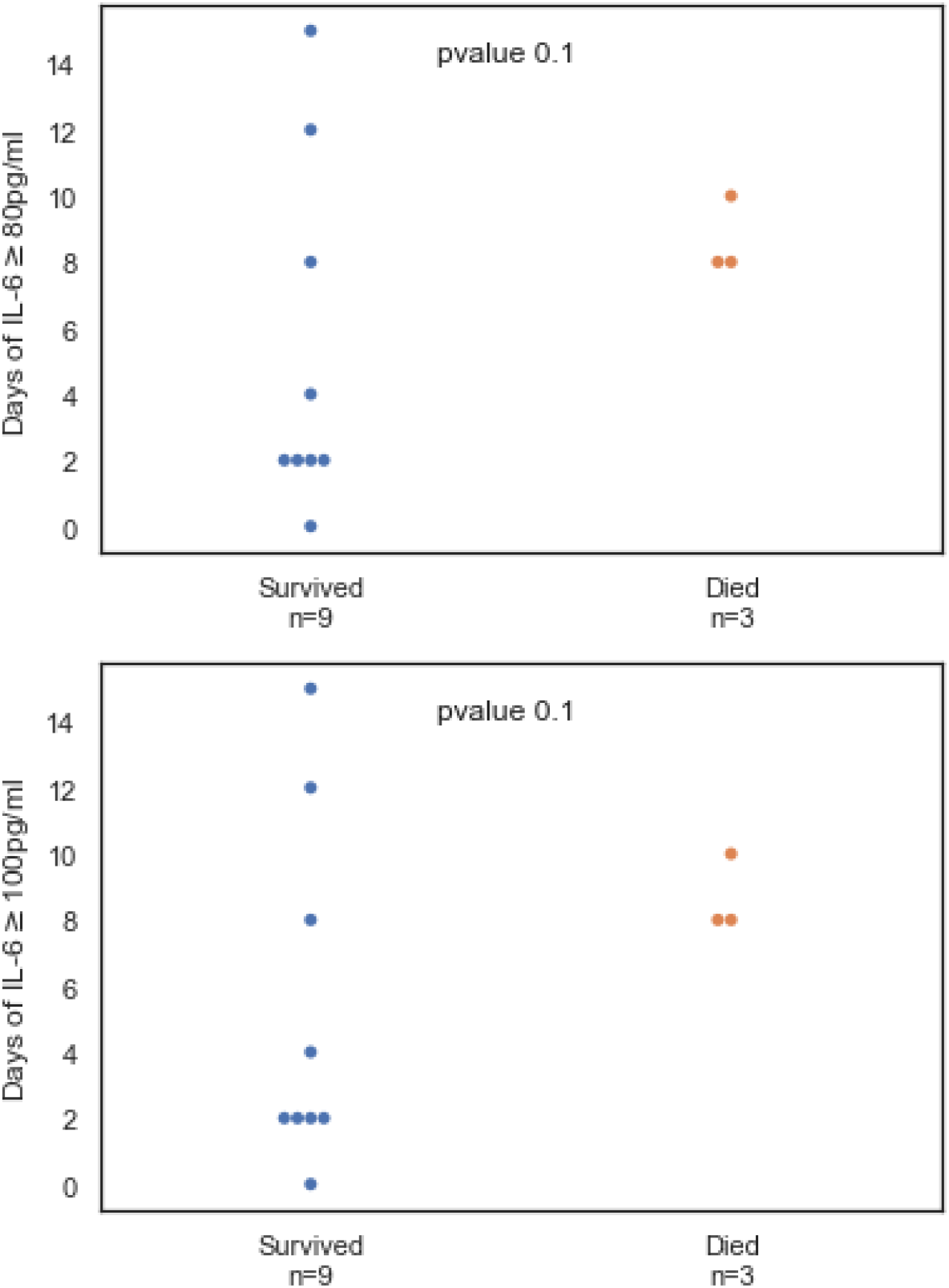
Mortality is not correlated with the number of days IL-6 > 80 pg/ml (upper panel) or number of days IL-6 > 100 pg/ml (lower panel) in the ICU patients (n = 12) IL-6 was measured at multiple time points during the 12 patient’s stay in the ICU. At least one IL-6 > 80 pg/ml (upper panel) or IL-6 > 100 pg/ml in a given day was sufficient for the analysis. Each circle represents a patient; blue dots represent patients who survived (n = 9) and red dots represent patients who died (n = 3).

